# Longitudinal decline in striatal DAT binding in LRRK2 Parkinson’s disease: connections with CSF α-Synuclein Seeding Activity

**DOI:** 10.1101/2025.05.19.25327958

**Authors:** Jing Wang, Xixi Sun, Yunfei Yin, Ruihua Cao

**Author notes:** **Correspondences:** Dr. Ruihua Cao.

## Abstract

**Objective:** Parkinson’s disease (PD) associated with mutations in the LRRK2 gene exhibits considerable pathological heterogeneity and may not present with Lewy body pathology. The α-Syn seed amplification assay (SAA) performed on cerebrospinal fluid (CSF) has emerged as a reliable *in vivo* biomarker of α-Syn aggregation. In this study, we aim to investigate the longitudinal trajectories of striatal dopaminergic imaging in LRRK2 PD patients stratified by CSF α-Syn SAA status.

**Methods:** Data were obtained from the Parkinson’s Progression Markers Initiative. CSF α-Syn aggregation was assessed using SAA. Striatal DAT specific binding ratios (SBR) were quantified using [¹²³I] FP-CIT SPECT at baseline, year 2, and year 4.

**Results:** At baseline, the α-Syn SAA-negative LRRK2 PD group exhibited higher DAT binding in the contralateral putamen and ipsilateral putamen compared to the SAA-positive group with comparable disease duration. Longitudinally, linear mixed-effects models demonstrated that the α-Syn SAA-negative LRRK2 PD maintained significantly higher DAT binding in both the contralateral and ipsilateral putamen over time. A significant group × time interaction was identified in the contralateral caudate, suggesting a slower rate of DAT loss in the α-Syn SAA-negative group. Sensitivity analyses restricted to participants with complete baseline and follow-up imaging data largely confirmed the main LMEM findings.

**Conclusions:** The observed differences in striatal dopaminergic degeneration between LRRK2 PD patients with and without detectable CSF α-synuclein aggregates may reflect region-specific vulnerability to underlying pathological processes. Our findings support the utility of CSF α-Syn SAA status as both a diagnostic and prognostic biomarker in LRRK2 PD.

## 1. Introduction

Parkinson’s disease (PD) is a progressive neurodegenerative disorder characterized by the loss of dopaminergic neurons in the substantia nigra and the accumulation of α-synuclein (α-Syn) aggregates in the form of Lewy bodies and Lewy neurites [4, 40]. However, in patients with PD carrying *LRRK2* mutations (LRRK2 PD), the underlying histopathology is remarkably heterogeneous. While a majority (60-80%) of LRRK2-related cases exhibit α-Syn positive Lewy body pathology, a considerable subset lacks detectable α-Syn aggregates [21, 36]. In these cases, the underlying pathology may instead involve isolated nigral degeneration or the deposition of other proteins like tau or TDP-43 [42, 43]. This pathological heterogeneity highlights the need for in vivo, pathology-driven biomarkers capable of distinguishing among these diverse molecular subtypes.

The α-Syn seed amplification assay (SAA) has emerged as a promising in vivo biomarker for detecting misfolded α-Syn in cerebrospinal fluid (CSF) with high sensitivity and specificity for Lewy body diseases such as PD and dementia with Lewy bodies (DLB) [13, 22, 33, 35, 37]. SAA detects α-Syn aggregation in CSF in a significant number of LRRK2 PD patients, but less frequently than in idiopathic PD (40% vs. 90%) [16]. Within the Parkinson’s Progression Markers Initiative (PPMI)—the largest available dataset for α-Syn SAA across genetically defined PD cohorts—approximately 67.5% of LRRK2 PD patients test positive for CSF α-Syn aggregation [6, 38]. Notably, clinical and biomarker differences have been observed between SAA-positive and SAA-negative LRRK2 PD individuals. Patients lacking CSF α-Syn aggregates tend to be older, female, have less olfactory impairment, exhibit milder motor symptoms and slower motor decline, and show higher serum neurofilament light chain (NfL) levels. Autopsy-CSF correlation analysis demonstrate that CSF SAA can accurately discriminate between LRRK2 PD cases with and without Lewy-type pathology [15, 18]. These findings suggest that SAA could serve as a valuable tool for stratifying patients in clinical trials targeting this genetic PD subtype.

[^123^I] FP-CIT single-photon emission computed tomography (SPECT) is a neuroimaging technique that quantifies striatal dopamine transporter (DAT) density—a surrogate marker of dopaminergic neurodegeneration [3, 12]. Given its utility in objectively tracking disease progression, we leveraged the PPMI dataset to investigate longitudinal changes in DAT imaging over four years in LRRK2 PD patients, stratified by their CSF α-Syn SAA status. By examining differences in dopaminergic decline between SAA-positive and SAA-negative LRRK2 PD patients, this study aims to enhance our understanding of how CSF α-Syn seeding activity influences dopaminergic neurodegeneration and clinical prognosis in this genetically defined PD subtype.

## 2. Materials and methods

### 2.1. Participants

Data were obtained from the PPMI database (www.ppmi-info.org/data), a multicenter, prospective cohort study. PPMI methods have been described elsewhere in detail [29, 32]. The PPMI study is registered with ClinicalTrials.gov (number NCT01141023). The LRRK2 PD cohort enrolled male or female participants aged 18 or older who met established diagnostic criteria for PD [1], had a disease duration of less than 7 years at screening, Hoehn and Yahr stage less than 4, and a confirmed LRRK2 mutation determined by the genetic core. The data used for this article were downloaded December 11, 2024.

Inclusion criteria for this analysis were as follows: (i) availability of α-Syn SAA result (see methods below) and (ii) available DAT imaging at baseline; for longitudinal analysis, participants were required to have at least two DAT imaging time points. Exclusion criteria include: (i) lowest putamen DAT specific binding ratio (SBR) ≥65% of the age- and sex-expected value in individuals who had a negative α-Syn SAA result, (ii) presence of known pathogenic GBA1 variant, and (iii)SAA results consistent with a multiple system atrophy-like (MSA-like) pattern [7].

### 2.2. DAT imaging

DAT-SPECT was performed as previously described [39]. ^123^I-FP-CIT SPECT imaging was conducted at participating PPMI sites and the acquired images were sent to the Institute of Neurodegenerative Disorders (IND, New Haven, CT, United States) for quality control and data extraction. SPECT image volumes were spatially normalized to standard Montreal Neurologic Institute space using the PMOD software (PMOD Technologies, Zurich, Switzerland). Next, the transaxial slice with the highest striatal uptake was identified and the eight hottest striatal slices around it were averaged to generate a single slice image. Regions of interest were then placed on the bilateral caudate, putamen, and the occipital cortex (reference tissue). SBRs were calculated for each of the four striatal regions (count density in the target region / count density in the reference region -1). The contralateral side was defined as the hemisphere opposite to the more clinically affected side of the body, whereas the ipsilateral side corresponded to the same side as the more affected body side.

For participants in the genetic cohorts, DAT imaging was performed at three visits: screening (baseline), year 2 and year 4. The percentage of age- and sex-expected lowest putamen SBR was determined using normative data derived from healthy controls in PPMI. A value of less than 65% of the expected binding in the lowest putamen was considered indicative of a quantitatively abnormal DAT-SPECT result.

### 2.3. Clinical Assessments

The clinical assessment battery included the Movement Disorders Society-Unified Parkinson’s Disease Rating Scale (MDS-UPDRS), which was assessed in both the medication “on” and “off” states after the initiation of dopaminergic therapy; for the purpose of this analysis, we utilized MDS-UPDRS part III in the “off” state. Other assessments include the Montreal Cognitive Assessment (MoCA) for global cognitive function, the 15-item Geriatric Depression Scale, the Scale for Outcomes for PD-autonomic function (SCOPA-AUT), the State and Trait Anxiety Scale (STAI), the modified Schwab and England Activities of Daily Living Scale, the Questionnaire for Impulsive-Compulsive Disorders in Parkinson’s Disease (QUIP), the Epworth Sleepiness Scale (ESS), the Rapid Eye Movement Sleep Behavior Disorder Screening Questionnaire (RBDSQ), and the University of Pennsylvania Smell Identification Test (UPSIT; Sensonics, Philadelphia, PA, USA). Hyposmia is defined as a UPSIT score at or below the 15th percentile, based on internal population norms [5]. Other measures included basic demographic characteristics and the use of dopaminergic medications, quantified using the levodopa equivalent daily dose (LEDD).

### 2.4. α-Syn seed amplification assay

The presence of aggregated α-Syn in CSF collected at either the baseline visit or the first-year follow-up visit was assessed using α-Syn SAA. Two versions of the assay were used within the PPMI study: a 150-hour reaction time assay as described [7, 38] and a 24-hour reaction time assay. These two assays have comparable performance characteristics, with equivalent sensitivities of 95% and specificities of 97% and 95%, respectively [26]. The Fmax (maximum raw fluorescence signal per well), T50 (time to reach 50% of the Fmax) and time to reach a target relative fluorescent units (RFU) threshold were used to define positive, inconclusive, negative and (for the 24-hour reaction time) MSA-like as described on PPMI website.

### 2.5. Genetic Testing

Genetic testing for the LRRK2 gene was conducted either locally at the study site or centrally through a recruitment initiative, using laboratories certified under the Clinical Laboratory Improvement Amendments (CLIA) or equivalent accredited facilities. The participants enrolled in the LRRK2 PD cohort were notified of their genetic testing results and received genetic counseling by phone or in person by certified genetic counselors or qualified site personnel. Dual mutation carriers for both LRRK2 and GBA were excluded from this analysis.

### 2.6. Statistical Methods

Statistical analyses were performed using IBM SPSS 26.0 software. Baseline descriptive statistics were reported as median and interquartile range (IQR) for continuous variables due to data skewness, and as frequency and percentage for categorical variables. Group differences in baseline demographic and clinical features were assessed using two-sample Wilcoxon rank sum test, chi-square test or Fisher’s exact test, as appropriate. To account for potential confounders, regression models were constructed to test the association between clinical and demographic features (predictors) and α-Syn SAA status (outcome), adjusting for age and sex. Years of education were additionally included as a covariate in models evaluating cognitive performance. All statistical tests were two-sided and p values ≤ 0.05 were considered statistically significant. As all analyses were explorative, we did not correct for multiple testing.

Linear mixed-effects models (LMEMs) were employed for longitudinal analyses, as they are well suited for handling repeated measures data, allowing for the modeling of autocorrelation and accommodating missing data. A restricted maximum likelihood (REML) approach was employed, with fixed effects including group, time, and their interaction (group × time), and a random intercept for each subject. Age and sex were accounted for all models and years of education were additionally included as a covariate in models evaluating cognitive performance. To address the impact of missing data, a sensitivity analysis was conducted by repeating the LMEM in participants with complete follow-up data.

## 3. Results

### 3.1. Baseline demographic and clinical characteristics

The study included a total of 141 participants carrying LRRK2 mutations: G2019S (n = 121), R1441G (n = 15), G2019S/G (n = 2), R1441C (n = 1), I2020T (n = 1) and N1437H (n = 1).

Details on demographic and clinical PD characteristics of CSF α-Syn SAA-positive and -negative participants at baseline are shown in Table 1. Compared to the α-Syn SAA-positive group, the α-Syn SAA-negative LRRK2 PD group were more likely to be female (60.4% vs 40.9%, OR = 2.209, p = 0.028), older (median age 69.09 vs 61.25, p < 0.001), had more family history of PD (87.5% vs 57.0%, OR = 5.283, p < 0.001; for male patients, 100% vs 54.5%, p < 0.001) and had better olfaction (median UPSIT score 30.0 vs 23.0, adjusted p < 0.001). Moreover, they exhibited milder motor symptoms on examination (median MDS-UPDRS III score 16.0 vs 23.0, adjusted p = 0.012) and were receiving lower doses of dopaminergic therapy (median LEDD 200mg vs 499 mg, adjusted p < 0.001) than the α-Syn SAA-negative group despite similar disease duration. In contrast to their milder motor symptoms, the α-Syn SAA-negative LRRK2 PD group showed poorer global cognitive performance compared to the α-Syn SAA-positive LRRK2 PD group after adjusting for age, sex, and years of education (median Moca score 25.00 vs 27.00, adjusted p = 0.013).

Striatal DAT SBRs at baseline are also summarized in Table 1. The α-Syn SAA-negative LRRK2 PD group exhibited higher DAT binding in the contralateral putamen (median SBR 0.70 vs 0.58; adjusted p = 0.006) and ipsilateral putamen (median SBR 0.92 vs 0.74; adjusted p = 0.001) compared to the α-Syn SAA-positive LRRK2 PD group, indicating less dopaminergic neurodegeneration compared to the α-Syn SAA-positive group. No significant differences in caudate DAT binding levels were observed between groups at baseline.

### 3.2. Longitudinal analyses

A total of 104 participants underwent at least two DAT imaging assessments, of whom 60 (57.7%) participants completed all three timepoints. Consistent with baseline findings, LMEM revealed that the α-Syn SAA-negative LRRK2 PD group continued to demonstrate milder motor symptoms (MDS-UPDRS part III, p = 0.020), fewer motor complications (MDS-UPDRS part IV, p = 0.016), and lower dopaminergic medication use (LEDD, p < 0.001) over time compared to the α-Syn SAA-positive group. Moreover, a significant group × time interaction was observed for MDS-UPDRS part II (p = 0.002), indicating a slower progression of self-reported functional impairment in the α-Syn SAA-negative LRRK2 PD group. However, group differences in MoCA scores were not sustained in the LMEM analysis (see Table 2 for details).

LMEM also showed persistently higher DAT binding in the contralateral and ipsilateral putamen (p = 0.016 and 0.017, respectively) in the α-Syn SAA-negative LRRK2 PD group compared to the α-Syn SAA-positive LRRK2 PD group. Additionally, a significant group × time interaction was identified in the contralateral caudate (p = 0.022), suggesting a slower rate of DAT loss in this region in the α-Syn SAA-negative group compared to the α-Syn SAA-positive group.

### 3.3. Sensitivity analyses

To ensure the robustness of the findings, sensitivity analyses were conducted on a subset of participants with complete baseline and follow-up data (n = 60). These analyses largely confirmed the main LMEM findings, aside from the group effects on SCOPA-AUT total score was marginally significant (p = 0.047), suggesting that the α-Syn SAA-negative LRRK2 PD group may experience slightly fewer automatic symptoms than their α-Syn SAA-positive counterparts (for details see Table 2).

## 4. Discussion

This study employed a longitudinal design to investigate the association between striatal dopaminergic dysfunction and CSF α-Syn SAA in LRRK2 PD patients. Our findings demonstrate that SAA-positive status is associated with a distinct pattern of dopaminergic terminal loss compared to SAA-negative status, characterized by greater dopaminergic loss in the putamen and a subsequently accelerated decline in the caudate as the disease progresses. The divergent pattern of dopaminergic degeneration may help explain the observed association between SAA positivity and more severe motor symptoms as well as faster motor progression. These results add to the growing body of evidence suggesting that SAA negativity may reflect alternative pathological mechanisms, potentially involving non-synucleinopathy-related neurodegeneration in LRRK2 PD.

Consistent with previous findings from the PPMI cohort [6, 38], we found SAA-positive LRRK2 PD patients were predominantly male, exhibit more pronounced motor disabilities and olfactory impairment. In contrast, SAA-negative LRRK2 PD patients are more likely to be female and had a relatively older age at disease onset compared with their SAA-positive counterparts. These distinctions may reflect fundamentally different pathogenic mechanisms: the underlying pathology of SAA-positive LRRK2 PD appears to confer greater vulnerability in males, while that of SAA-negative LRRK2 PD may preferentially affect females and exhibit stronger age dependency. Age and sex have been reported to play a role in LRRK2 PD pathologies [14, 19, 41]. For example, an animal study reported that prodromal intestinal inflammation promotes the development of PD-like phenotypes in male—but not female—LRRK2 G2019S mice, accompanied by early accumulation of α-synuclein in gut-associated myeloid cells [14]. Additionally, the R1441C-LRRK2 mutation has been shown to induce myeloid immune cell exhaustion in an age- and sex-dependent manner in mice [41].

In the present study, we observed that SAA-negative LRRK2 PD patients were more likely to report family history of PD compared SAA-positive patients. Notably, this difference seemed specific to male patients: those in the SAA-positive group had a significantly higher prevalence of family history of PD compared to the SAA-negative group. Specifically, all male patients without a family history of PD (n = 25) were SAA-positive, while all SAA-negative patients without a family history (n = 6) were females. These findings suggest that biological factors unique to females—such as hormonal regulation, immune function, or metabolic processes—may interact with de novo or low-penetrance LRRK2 mutations to drive a distinct, SAA-negative disease process.

A previous study combining quantitative MRI techniques with DAT imaging reported that α-Syn aggregation was associated with reduced regional brain volumes, altered caudal functional connectivity. However, no significant differences in striatal SBRs were detected between PD-SAA+ and PD-SAA-groups. This lack of significance may be attributed to their limited sample size (n = 41) and the heterogeneity of the PD cohort, which included idiopathic PD, LRRK2 PD, and GBA PD [10]. Here, we utilized a larger and genetically homogeneous cohort of LRRK2 PD patients from the PPMI dataset. We found significant differences between SAA-positive and SAA-negative LRRK2 PD patients. FP-CIT SPECT studies in healthy volunteers have consistently reported an age-related decline in striatal SBRs, ranging between 2.5% and 9.6% per decade [23]. In our study, SAA-positive patients exhibited more severe DAT loss despite being younger than SAA-negative patients. This suggests that pathological damage to the dopaminergic system is more pronounced in SAA-positive individuals.

The loss of dopaminergic neurons in the substantia nigra pars compacta does not uniformly affect all parts of the striatum. Dopaminergic projections to the putamen are generally affected earlier and more significantly than those to the caudate nucleus [25]. In our study, a value of less than 65% of the expected binding in the lowest putamen was considered indicative of a quantitatively abnormal DAT-SPECT result. Both SAA-positive and SAA-negative LRRK2 PD patients showed DAT loss in the putamen, but the loss was less prominent in SAA-negative patients from baseline through the 4-year follow-up. As the disease progresses, DAT reduction rates were faster in the contralateral caudate in SAA-positive patients compared to SAA-negative patients. Our data suggested that striatal dopaminergic neurons are more susceptible to the intrinsic pathology underlying SAA positivity. The distinct pattern of dopaminergic degeneration observed may help explain the association between SAA positivity and greater symptom severity and faster motor progression [9, 12, 30],

In our study, contrary to motor symptoms, the SAA-negative group showed more pronounced global cognitive dysfunction at baseline, as assessed by the MoCA. These results should be interpreted with caution, given differences in age, sex and education differences between the two groups. Furthermore, the observed group differences were not sustained in the longitudinal analyses. Nevertheless, the results raise the possibility that SAA-negative *LRRK2* PD patients may be at increased risk for cognitive decline. A previous PPMI study reported elevated total tau and phosphorylated tau levels in the SAA-negative group; however, these differences did not remain statistically significant when adjusting for age [6]. There is a possibility that SAA-negative cases may be more susceptible to tauopathy-driven neurodegeneration, which could preferentially affect cortical structures and contribute to more severe cognitive impairment. Future studies with more rigorous control of potential confounders are warranted to further compare cognitive function between SAA-positive and SAA-negative groups. In particular, multimodal imaging approaches will be critical for elucidating the structural and functional differences underlying these divergent clinical profiles.

Consistent with previous autopsy-CSF correlation analysis demonstrating that CSF α-Syn SAA can discriminate, with high sensitivity and specificity, LRRK2 PD cases with Lewy-type pathology from those without [15, 18], in all 27 cases that came to post-mortem examination in PPMI participants, all participants with positive α-Syn SAA results (n = 21, 2 of them were LRRK2 PD cases) had typical Lewy pathology, whereas the 2 case with negative α-Syn SAA result (an LRRK2 PD case and an idiopathic PD case) had no Lewy pathology. LRRK2 is a critical regulator of multiple cellular processes, including autophagy [2, 31], apoptosis [24], mitochondrial function [28] and neuroinflammation [34]. Dysregulation of LRRK2 function has been shown to interact with both α-synuclein and tau pathologies through these cellular pathways [8, 17, 20, 27]. Given the heterogenous pathological landscape observed in LRRK2 PD, it remains unclear whether and how LRRK2 activity might impact the development of protein aggregate pathologies [11]. Further research is needed to elucidate the mechanistic links between LRRK2 dysfunction and distinct neurodegenerative proteinopathies.

There are some limitations in our study. First, the sample size remains relatively modest and we were unable to perform separate analyses for each mutation type. Nonetheless, to our knowledge, this represents the largest comprehensive study of LRRK2 PD to data that incorporates CSF α-Syn SAA data alongside detailed clinical and DAT imaging assessments. As the PPMI is continuously expanding, future data releases are expected to increase the sample size, thereby allowing for mutation-specific analyses and further validation of our findings. Second, the presence of missing data within the longitudinal cohort poses challenges for statistical inference and interpretation. However, the use of linear mixed-effects models is appropriate for such longitudinal designs, as these models account for both repeated measurements and missingness. In addition, we conducted sensitivity analyses to assess the robustness of our results.

In conclusion, our study identified several distinguishing clinical and striatal DAT imaging patterns between LRRK2 PD cases with and without detectable α-Syn aggregates in the CSF, as determined by SAA. Our findings show that α-Syn SAA positivity is associated with greater dopaminergic loss in the putamen and a more rapid decline in dopaminergic integrity in the caudate. This distinct pattern of striatal dopaminergic degeneration may reflect differential regional vulnerability to underlying pathological processes in the two groups, potentially driven by the presence or absence of α-synuclein pathology. These results support the potential of CSF α-Syn SAA status as both a diagnostic and prognostic biomarker within the LRRK2 PD population.

## Author contributions

J.W. and R.C. conceptualized and designed the study. X.S. and Y.Y. performed data processing and statistical analyses. J.W. and R.C. interpreted the results. J.W. drafted the initial manuscript. All authors critically reviewed and edited the manuscript and approved the final version.

## Funding

This work was supported in part by the National Natural Science Foundation of China [grant numbers 82401745, 82471271]. The Parkinson’s Progression Markers Initiative (PPMI)—a public–private partnership—is funded by the Michael J. Fox Foundation for Parkinson’s Research and funding partners, including 4D Pharma, Abbvie, AcureX, Allergan, Amathus Therapeutics, Aligning Science Across Parkinson’s, AskBio, Avid Radiopharmaceuticals, Bial Portela, BioArctic, Biogen, Biohaven, BioLegend, BlueRock Therapeutics, Bristol-Myers Squibb, Calico Labs, Capsida Biotherapeutics, Celgene, Cerevel Therapeutics, Coave Therapeutics, DaCapo Brainscience, Denali, Edmond J. Safra Foundation, Eli Lilly, Gain Therapeutics, General Electric HealthCare, Genentech, GlaxoSmithKline (GSK), Golub Capital, Handl Therapeutics, Insitro, Jazz Pharmaceuticals, Johnson & Johnson Innovative Medicine, Lundbeck, Merck, Meso Scale Discovery, Mission Therapeutics, Neurocrine Biosciences, Neuron23, Neuropore Therapies, Pfizer, Piramal, Prevail Therapeutics, Roche, Sanofi, Servier, Sun Pharma Advanced Research Company, Takeda, Teva, Union Chimique Belge (UCB), Vanqua Bio, Verily, Voyager Therapeutics, the Weston Family Foundation and Yumanity Therapeutics.

## Data Availability

All data produced are available online at https://www.ppmi-info.org/.

## Ethics declarations

## Conflict of interest

The authors declare that they have no conflict of interest.

## Ethical approval

The study was approved by the institutional review board at each site, and participants provided written informed consent. This research was conducted ethically in accordance with the Declaration of Helsinki.

## Notes

### Competing Interest Statement

The authors have declared no competing interest.

### Author Declarations

The study used (or will use) ONLY openly available human data that were originally located at https://www.ppmi-info.org/. The study was approved by the institutional review board at each site, and participants provided written informed consent.

