## Supplementary material for "Longitudinal decline in striatal DAT binding in LRRK2 Parkinson’s disease: connections with CSF α-Synuclein Seeding Activity": Table1

Tabel 1 Baseline demographics, PD characteristics and striatal DAT SBRs

| **Variable** | **SSA negative**  **(n = 48)** | **SSA positive**  **(n = 93)** | ***p*** | **adj. *p*** |
| --- | --- | --- | --- | --- |
| **Demographics** |  |  |  |  |
| Age | 69.09 [65.31, 72.11] | 61.25 [55.04, 66.78] | **<0.001** |  |
| Age of onset | 64.57 [58.72, 68.63] | 57.23 [47.67, 61.98] | **<0.001** |  |
| Sex, male, n (%) | 19 (39.6) | 55 (59.1) | **0.028** |  |
| Education, y | 14.50 [10.00, 17.00] | 17.00 [14.00, 19.00] | **0.001** |  |
| Family history of PD |  |  | **<0.001** |  |
| First/second degree, n (%) | 42 (87.5) | 53 (57.0) |  |  |
| Male (n) | 19 | 30 | **<0.001** |  |
| None, n (%) | 6 (12.5) | 40 (43.0) |  |  |
| Male (n) | 0 | 25 |  |  |
| **PD characteristics** |  |  |  |  |
| Duration, months | 23.30 [10.58, 49.88] | 24.80 [13.33, 51.07] | 0.643 |  |
| Hoehn & Yahr, off | 2.00 [1.50, 2.00] | 2.00 [1.00, 2.00] | 0.331 | 0.981 |
| Modified Schwab & England ADL | 90 [90, 100] | 90 [90, 100] | 0.525 | 0.175 |
| MDS-UPDRS part I | 6.50 [2.50, 10.00] | 7.00 [3.50, 11.00] | 0.703 | 0.925 |
| MDS-UPDRS part II | 5.50 [2.00, 8.50] | 7.00 [3.00, 10.00] | 0.170 | 0.415 |
| MDS-UPDRS part III, off | 16.00 [12.50, 23.50] | 23.00 [14.00, 30.00] | **0.018** | **0.012** |
| MDS-UPDRS part IV | 0.00 [0.00, 0.00] | 0.00 [0.00, 4.00] | **0.023** | 0.226 |
| TD/PIGD classification, off  non_TD |  |  | 0.670 | 0.601 |
| TD | 22 (45.8) | 40 (43.0) |  |  |
| non-TD (PIGD or Indeterminate) | 22 (45.8) | 34 (36.6) |  |  |
| Unknown | 4 (8.3) | 19 (20.4) |  |  |
| LEDD, mg | 200.0 [100.0, 356.2] | 499.0 [300.0, 810.0] | **<0.001** | **<0.001** |
| UPSIT raw score | 30.0 [26.0, 34.0] | 23.0 [17.00, 29.5] | **<0.001** | **<0.001** |
| UPSIT age-/sex- adjusted percentile |  |  |  |  |
| Normosmia (>15%), n (%) | 34 (70.8) | 20 (21.5) | **<0.001** |  |
| Hyposmia (<= 15%), n (%) | 13 (27.1) | 68 (73.1) |  |  |
| Unknown, n (%) | 1 (2.1) | 5 (5.4) |  |  |
| ESS | 5.50 [4.00, 9.00] | 7.00 [4.00, 11.00] | 0.127 | 0.773 |
| RBDSQ | 3.00 [1.50, 4.00] | 4.00 [2.00, 5.00] | 0.130 | 0.273 |
| SCOPA-AUT total score | 11.00 [6.00, 16.50] | 10.00 [6.50, 16.00] | 0.657 | 0.766 |
| MoCA | 25.00 [22.50, 27.00] | 27.00 [25.50, 29.00] | **<0.001** | **0.013** |
| GDS | 2.00 [1.00, 5.00] | 2.00 [0.00, 4.00] | 0.159 | 0.461 |
| STAI state | 34.00 [26.50, 42.00] | 33.00 [24.00, 42.00] | 0.780 | 0.619 |
| STAI trait | 36.00 [28.50, 44.00] | 34.00 [27.00, 41.00] | 0.344 | 0.245 |
| QUIP score | 0.00 [0.00, 1.00] | 0.00 [0.00, 1.00] | 0.671 | 0.923 |
| Any QUIP disorder, n (%) | 16 (33.3) | 35 (37.6) | 0.614 | 0.737 |
| **DAT SBRs** |  |  |  |  |
| Contralateral caudate | 1.68 [1.38, 1.91] | 1.66 [1.31, 2.06] | 0.932 | 0.784 |
| Ipsilateral caudate | 1.95 [1.57, 2.27] | 1.93 [1.51, 2.35] | 0.997 | 0.810 |
| Contralateral putamen | 0.70 [0.57, 0.89] | 0.58 [0.48, 0.76] | **0.003** | **0.006** |
| Ipsilateral putamen | 0.92 [0.68, 1.09] | 0.74 [0.57, 0.92] | **0.008** | **0.001** |

Note: Descriptive statistics were used to represent the baseline features of participants, with continuous variables expressed as median and interquartile range (IQR) and categorical variables expressed as frequency and percentage. We conducted the two-sample Wilcoxon rank sum test, chi-square test or Fisher’s exact test to compare baseline demographic data of different subgroups. Linear regression and logistic regression models using α-Syn SAA as predictor of outcome and adjusting for age, sex and years of education (only for MoCA score) were used for continuous and categorical variables, respectively.

Abbreviations: ADL, Activity of Daily Living Scale; DAT, Dopamine transporter; ESS, Epworth Sleepiness Scale; GDS, Geriatric Depression Scale; LEDD, Levodopa Equivalent Daily Dose; MDS-UPDRS, Movement Disorder Society-Sponsored Revision Unified Parkinson's Disease Rating Scale; MoCA, Montreal Cognitive Assessment; PD, Parkinson's disease; PIGD, Postural Instability and Gait Difficulty; QUIP, Questionnaire for Impulsive-Compulsive Disorders in Parkinson’s Disease; RBDSQ, Rapid-eye-movement sleep Behavior Disorder Screening Questionnaire; SAA, Seed Amplification Assay; SBR, Specific binding ratios; SCOPA-AUT, Scales for Outcomes in Parkinson's disease – Autonomic; STAI: State–Trait Anxiety Inventory; TD, Tremor-Dominant; UPSIT, University of Pennsylvania Smell Identification Test.
