## Supplementary material for "Longitudinal decline in striatal DAT binding in LRRK2 Parkinson’s disease: connections with CSF α-Synuclein Seeding Activity": Table2

Table 2 Linear Mixed-Effects Models of clinical and DAT imaging trajectories

| **Variable** | **Main analysis (n = 104)** | | **Sensitivity analysis (n = 60)** | |
| --- | --- | --- | --- | --- |
|  | **Group effect** | **Group x Time effect** | **Group effect** | **Group x Time effect** |
| **PD characteristics** |  |  |  |  |
| Hoehn & Yahr, off | 0.680 | 0.156 | 0.861 | 0.072 |
| Modified Schwab & England ADL | 0.209 | 0.344 | 0.178 | 0.761 |
| MDS-UPDRS part I | 0.948 | 0.382 | 0.456 | 0.326 |
| MDS-UPDRS part II | 0.142 | **0.002** | 0.093 | **0.009** |
| MDS-UPDRS part III, off | **0.020** | 0.219 | **0.004** | 0.103 |
| MDS-UPDRS part IV | **0.016** | 0.107 | **0.012** | 0.316 |
| LEDD, mg | **<0.001** | 0.436 | **0.010** | 0.848 |
| MoCA | 0.111 | 0.297 | 0.594 | 0.629 |
| QUIP score | 0.438 | 0.424 | 0.247 | 0.852 |
| ESS | 0.637 | 0.074 | 0.173 | 0.377 |
| RBDSQ | 0.074 | 0.746 | 0.106 | 0.745 |
| GDS | 0.878 | 0.394 | 0.804 | 0.411 |
| STAI state | 0.676 | 0.534 | 0.866 | 0.678 |
| STAI trait | 0.753 | 0.907 | 0.948 | 0.243 |
| SCOPA-AUT total score | 0.426 | 0.072 | **0.047** | 0.094 |
| **DAT SBRs** |  |  |  |  |
| Contralateral caudate | 0.978 | **0.022** | 0.698 | **0.004** |
| Ipsilateral caudate | 0.814 | 0.296 | 0.285 | 0.583 |
| Contralateral putamen | **0.016** | 0.688 | **0.028** | 0.996 |
| Ipsilateral putamen | **0.017** | 0.925 | **0.028** | 0.974 |

Note: A restricted maximum likelihood (REML) approach was employed, with fixed effects including group, time, and their interaction (group×time), and a random intercept for each subject. Age and sex were accounted for all models and years of education were additionally included as a covariate in models evaluating cognitive performance. To address the impact of missing data, a sensitivity analysis was conducted by repeating the LMEM in participants with complete follow-up data.

Abbreviations: ADL, Activity of Daily Living Scale; DAT, Dopamine transporter; ESS, Epworth Sleepiness Scale; GDS, Geriatric Depression Scale; LEDD, Levodopa Equivalent Daily Dose; MDS-UPDRS, Movement Disorder Society-Sponsored Revision Unified Parkinson's Disease Rating Scale; MoCA, Montreal Cognitive Assessment; PD, Parkinson's disease; QUIP, Questionnaire for Impulsive-Compulsive Disorders in Parkinson’s Disease; RBDSQ, Rapid-eye-movement sleep Behavior Disorder Screening Questionnaire; SBR, Specific binding ratios; SCOPA-AUT, Scales for Outcomes in Parkinson's disease – Autonomic; STAI: State–Trait Anxiety Inventory.
